# Coupling Wastewater-Based Epidemiological Surveillance and Modelling of SARS-COV-2/COVID-19: Practical Applications at the Public Health Agency of Canada

**DOI:** 10.1101/2022.06.26.22276912

**Authors:** Meong Jin Joung, Chand S Mangat, Edgard Mejia, Audra Nagasawa, Anil Nichani, Carol Perez-Iratxeta, Shelley W Peterson, David Champredon

## Abstract

Wastewater-based surveillance (WBS) of SARS-CoV-2 offers a complementary tool for clinical surveillance to detect and monitor Coronavirus Disease 2019 (COVID-19). Since both symptomatic and asymptomatic individuals infected with SARS-CoV-2 can shed the virus through the fecal route, WBS has the potential to measure community prevalence of COVID-19 without restrictions from healthcare-seeking behaviors and clinical testing capacity. During the Omicron wave, the limited capacity of clinical testing to identify COVID-19 cases in many jurisdictions highlighted the utility of WBS to estimate disease prevalence and inform public health strategies. However, there is a plethora of in-sewage, environmental and laboratory factors that can influence WBS outputs. The implementation of WBS therefore requires a comprehensive framework to outline an analysis pipeline that accounts for these complex and nuanced factors. This article reviews the framework of the national WBS conducted at the Public Health Agency of Canada to present WBS methods used in Canada to track and monitor SARS-CoV-2. In particular, we focus on five Canadian cities – Vancouver, Edmonton, Toronto, Montreal and Halifax – whose wastewater signals are analyzed by a mathematical model to provide case forecasts and reproduction number estimates. This work provides insights on approaches to implement WBS at the national scale in an accurate and efficient manner. Importantly, the national WBS system has implications beyond COVID-19, as a similar framework can be applied to monitor other infectious disease pathogens or antimicrobial resistance in the community.

## Introduction

Epidemics caused by infectious pathogens are traditionally monitored through clinical surveillance of individuals. Wastewater-based surveillance (WBS) is an alternative epidemiological surveillance approach that consists of assessing the concentration of a pathogen of interest in wastewater to estimate its associated infection prevalence in a community. WBS has been integrated as part of poliovirus eradication initiatives since 2010 (HOVI et al., 2012). In Canada, it has been used to monitor drug consumption in multiple cities since 2019 (Yargeau, Taylor, Li, Rodayan, & Metcalfe, 2014, Statistics Canada, 2022). During the COVID-19 pandemic, WBS has attracted a lot of attention for surveillance of SARS-CoV-2 (the virus causing COVID-19) both in Canada and globally (Naughton et al., 2021). WBS provides a complementary tool to the clinical surveillance to detect and monitor trends of disease caused by SARS-CoV-2. As opposed to clinical surveillance of COVID-19 (Hilborne, Wagner, Cabreros, & Brook, 2020), WBS is not limited by under-diagnosis of asymptomatic individuals because most individuals infected with SARS-CoV-2 shed viral particles in their stools (Miura, Kitajima, & Omori, 2021; Sangkham, 2021). WBS utilizes a pooled community sample from the catchment area of a sampling location to measure the viral concentration of SARS-CoV-2 within the community (Shah et al., 2022). Multiple studies have shown that SARS-CoV-2 viral concentration measured in wastewater correlates with the real prevalence affecting the community living in the catchment area (Bonanno Ferraro et al., 2021; Medema, Heijnen, Elsinga, Italiaander, & Brouwer, 2020).

WBS has garnered high interest during the emergence of the variant of concern (VOC) Omicron in November 2021 (World Health Organization, 2021). Its large number of genetic mutations compared to the previous circulating lineages conferred the variant a higher transmissibility and immune escape that fueled its rapid growth (Khandia et al., 2022). Hence, during the Omicron wave, testing capacities in many countries, including in major Canadian cities, were overwhelmed, forcing the Polymerase Chain Reaction (PCR)-testing of SARS-CoV-2 in clinical samples to be restricted to certain high-risk or vulnerable populations (Public Health Ontario, 2021; World Health Organization, 2022). Previous research has demonstrated that SARS-CoV-2 was detected in 29-100% of fecal samples in infected individuals (Kitajima et al., 2020). In addition, Shah *et al*. reviewed that WBS detection of SARS-CoV-2 preceded confirmed clinical cases by up to 5 to 63 days (Shah et al., 2022). The Omicron wave demonstrated WBS can be used as an alternative measure of disease prevalence when clinical surveillance is limited by overwhelming demand or test-seeking behaviours and may serve as an early indicator of COVID-19 presence to inform testing and public health strategies at the community level (Wu et al., 2022). Overall, WBS offers a non-invasive and low-cost method to estimate the community prevalence of COVID-19 that complements the limitations of the traditional clinical surveillance.

However, WBS is not free of biases and uncertainties. WBS can be influenced by various pre-and post-analytical factors including methods of sample collection and storage, laboratory analysis protocol, engineering of the sewer network and wastewater treatment plants (WWTP), changes in weather conditions and data analysis procedures (Bertels et al., 2022; McCall et al., 2021; Wade et al., 2022). Moreover, since WBS for SARS-CoV-2 is still in its infancy, there is a lack of standardized procedures to address these factors. Considering the potential sensitivity of WBS data to these factors, it is crucial to establish an analysis pipeline that specifies standardized protocols and methodologies from sample collection to analysis to ensure the accuracy of WBS. While it may be impossible to control for some sources of uncertainty, minimizing their effects remains crucial. Importantly, there is a need to implement a framework to combine the results of WBS and clinical surveillance to clearly communicate the epidemiological findings to inform public health strategies (Smith et al., 2021).

In Canada, WBS is performed by laboratories at federal, provincial and municipal levels as well as academic groups (Naughton et al., 2021). The National Microbiology Laboratory (NML) at the Public Health Agency of Canada (PHAC) collates and analyzes samples from multiple provinces to conduct WBS at the national level. The objective of this review paper is to provide a comprehensive overview of the analysis pipeline of WBS at the NML and a framework to incorporate WBS and clinical surveillance to enhance the national surveillance of COVID-19. We describe how mathematical modelling can be utilized to facilitate the interpretation of WBS outputs. In addition, we assess key factors that influence WBS signals at each step of the pipeline and methods to address them to increase the usefulness of WBS data.

## Wastewater-based surveillance pipeline

The Canadian national WBS program involves the collaboration of municipal WWTP and multiple government divisions and agencies, including Statistics Canada, the NML and PHAC. A data pipeline was developed to streamline the WBS processes from sample collection to reporting in an accurate and timely manner (Figure 1).

**FIGURE 1.**
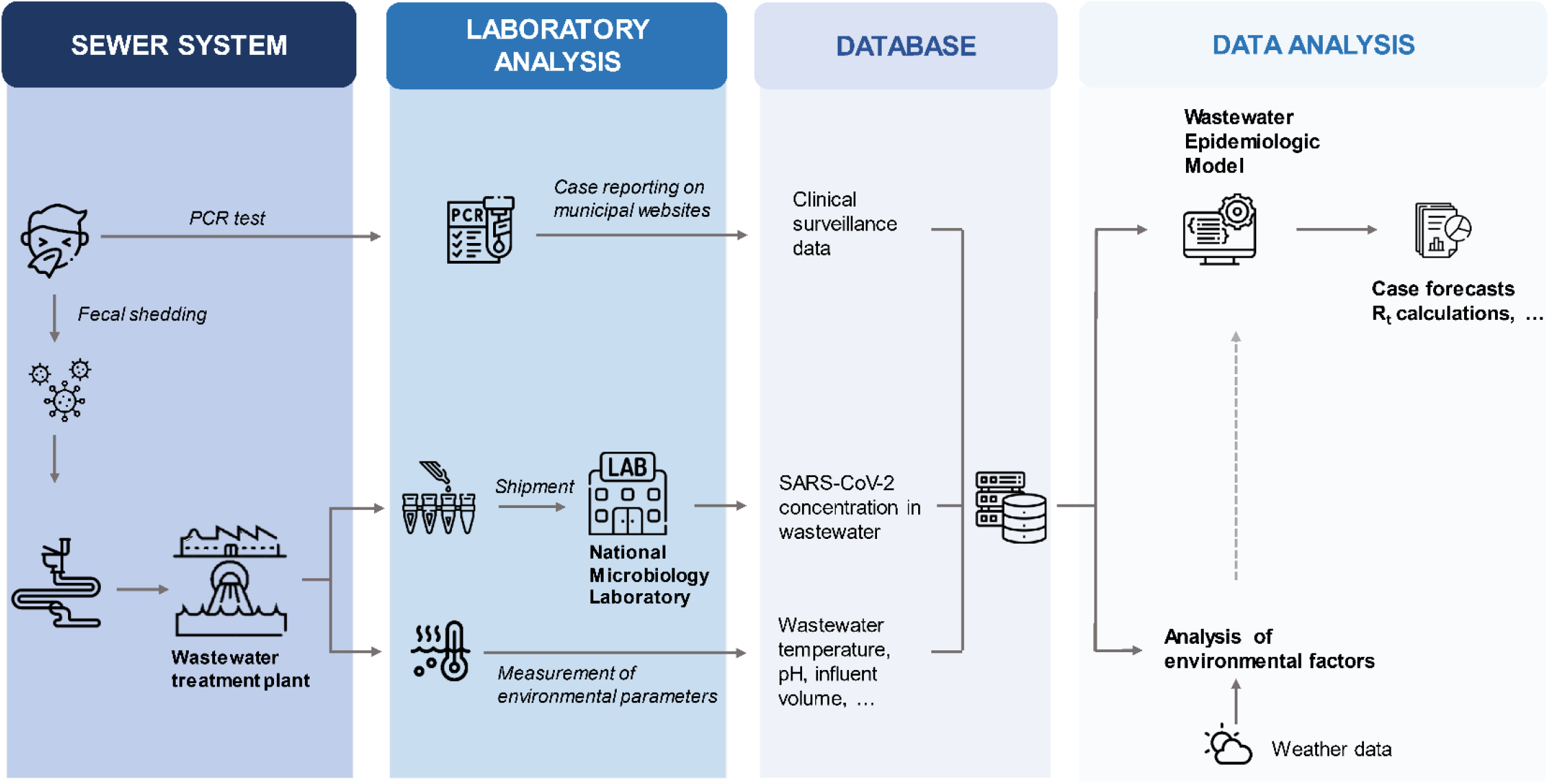
Data and analysis pipeline at PHAC/NML. While a fraction of clinical SARS-CoV-2 infections are asymptomatic and may be missed by traditional clinical surveillance, fecal shedding likely occurs in most infected individuals. These two types of events, symptomatic manifestations and fecal shedding, are at the origin of two data streams: clinical and wastewater. At PHAC/NML, wastewater is sampled at the treatment plant, typically covering a large population. RT-qPCR of clinical cases is usually performed on a multitude of clinical samples to assess the infection status for tested individuals, whereas the wastewater assesses the SARS-CoV-2 concentration in a single sample to provide a proxy for the community-level prevalence. While clinical surveillance may be biased and missed entire demographic groups, the wastewater signal can be affected by environmental variables independent of the epidemic (e.g., temperature, industrial waste, weather). Clinical and wastewater data streams can be linked using the postal code of reported cases and sewage maps of sampling locations. Data is saved in a database in a standardized format for various municipalities across Canada to provide scalability of data analysis. This database is then used to fit epidemic models to estimate unobserved variables (e.g., reproduction number R_t_) and perform short-term forecasts.

### Data collection

The Canadian Wastewater Survey (CWS) jointly led by Statistics Canada and PHAC currently involves 102 WWTP’s across Canada. We focus on 15 WWTP’s of five cities – Vancouver, Edmonton, Toronto, Montreal and Halifax – where mathematical modelling is applied to analyze the trends of SARS-CoV-2 (Figure 2). The wastewater sampling in the five cities began in September 2020. The samples are collected approximately twice a week from the raw influents. Samples are collected before de-gritting in one WWTP in Edmonton, three WWTPs in Montreal and three WWTPs in Vancouver and post-grit removal in four WWTPs in Toronto and two WWTPs in Vancouver. Wastewater can be sampled using composite or grab sample methods. Grab sampling constitutes rapid sampling at a specific point in time, which represents the influent at that time. Therefore, the results are more subject to changes in the influent flow of the day. Composite sampling involves collecting multiple samples using an automatic sampler during a set time-period (typically 24 hours) to represent the wastewater composition for that period. For the CWS, the composite sampling method was used where automatic samplers collected wastewater samples during a 24h period. These samples were temporarily stored and shipped at 4°C to the NML in Winnipeg, Manitoba.

**FIGURE 2.**
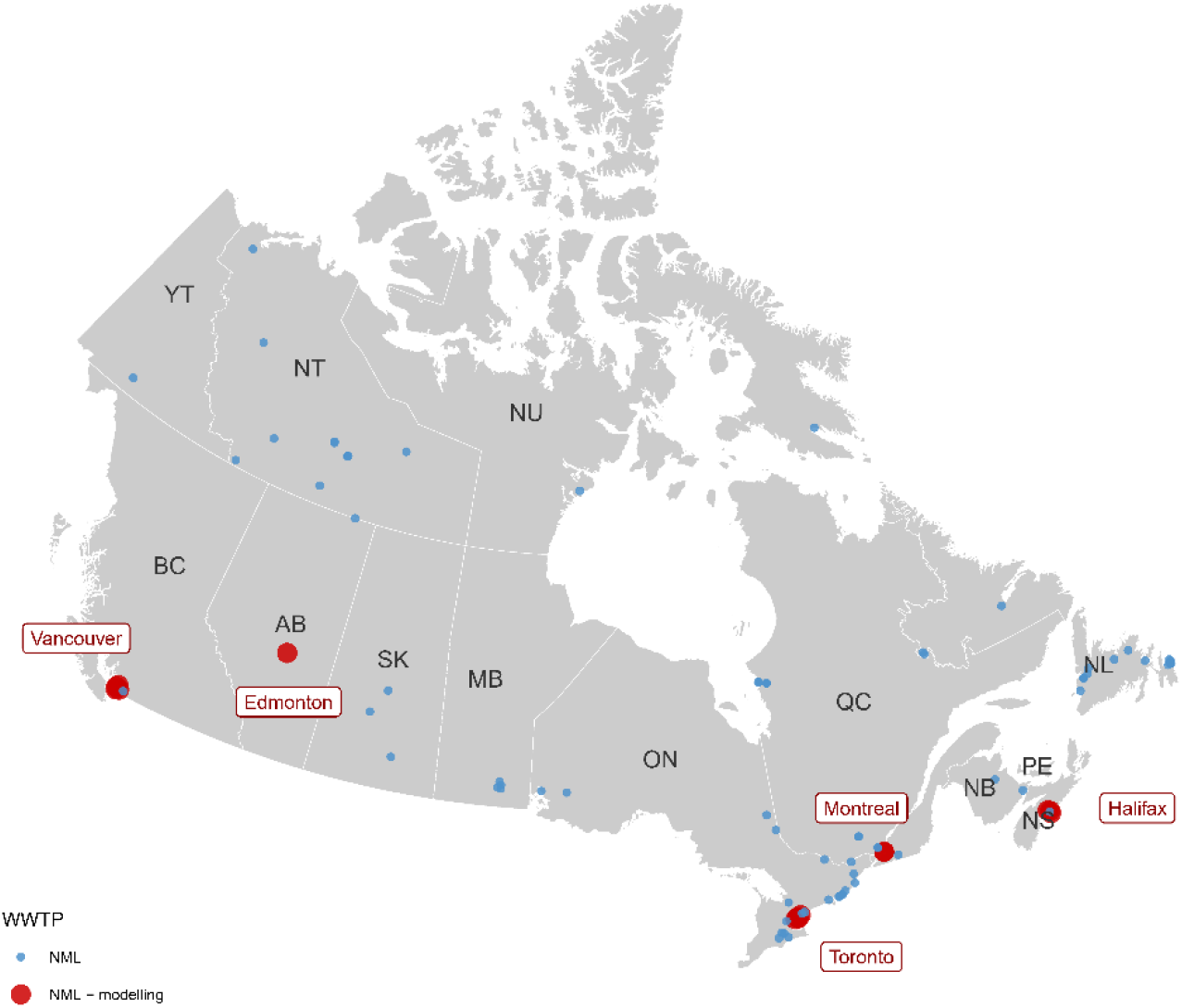
Wastewater-based surveillance (WBS) of COVID-19 is actively conducted by federal, provincial, and municipal governments and academic institutions. While the current framework utilizes data from five cities - Vancouver, Edmonton, Toronto, Montreal and Halifax - which are used for the Wastewater Epidemiologic Model to provide case forecasts, the data pipeline has the potential to expand to other cities and provide trends and/or forecasts of SARS-CoV-2 across Canada with the incorporation of concurrent WBS programs. The location of these WBS programs, conducted by the NML, academic institutions, and collaborators of NML, are plotted below to visualize the total coverage of COVID-19 WBS in Canada. Three WWTP sites that are monitored by NML are not included in the map due to privacy policies.

In addition to sample collection, wastewater quality and environmental parameters of the wastewater such as influent daily volume, temperature and pH are measured at the WWTP. The wastewater data from the NML are collated together with the environmental parameters from each WWTP by Statistics Canada for data management and shared with PHAC/NML.

Clinical surveillance data are retrieved from publicly available sources on municipal or provincial websites for each city. When available (e.g., Toronto, Vancouver), we collect data at the sub-municipal level to map the spatial location of the clinical reported cases with the catchment area of each WWTP. Weather-related environmental data, including amount of precipitation and snow on ground for each city, are obtained from Environment Canada (https://climate.weather.gc.ca/historical_data/search_historic_data_e.html).

### Laboratory analysis of SARS-CoV-2 concentration

SARS-CoV-2 concentration was measured with two methods. The laboratory protocols for the two methods were described in detail by Nourbakhsh *et al* (Nourbakhsh et al., 2022). Briefly, before February 12, 2021, SARS-CoV-2 RNA was extracted from the liquid supernatant portion of clarified wastewater samples. However, early studies found that the solid portion of clarified wastewater samples yield a higher viral concentration (Li et al., 2021, Palmer et al., 2021, Kim et al., 2022). Therefore, after February 12, 2021, RNA extraction was performed on the solid pellet after clarification. In Vancouver, the method based on liquid supernatant is still performed in addition to the solid pellet extraction. The change in protocol improved the efficiency of RNA quantification.

## Data analysis and modelling

### Data quality and sources of uncertainty

WBS data are influenced by several factors, including environmental conditions, laboratory protocols and engineering of the WWTP’s. We conducted investigations into these factors to account for them in our analyses.

#### Environmental factors

Environmental factors such as precipitation or snowmelts have been described as critical factors that could influence viral signals in the wastewater (Lazuka et al., 2021). However, the impact of environmental factors could vary depending on the type of the sewer system serviced by a WWTP. There are two major types of sewer systems – combined or sanitary. Combined systems (CS) collect storm water from surface runoff and wastewater together within the same pipes. While CS would only collect wastewater as influent water to the WWTP during dry weather, wet weather or high precipitation events (including snowmelts) would increase the influent flow rate and dilute viral concentration present in the wastewater (Lazuka et al., 2021). In contrast, sanitary systems mostly separate storm water and sewage, which means the influent volume do not significantly change based on the weather, avoiding the dilution of the viral signal.

CS are present in older parts of the cities monitored by PHAC. In order to ensure the quality of WBS, we investigated the potential mediating effects of precipitation on the WBS SARS-CoV-2 signal. Our analyses (manuscript in preparation) revealed that while some fluctuations in influent volume were recorded with changes in precipitation, they do not appear to significantly impact the SARS-CoV-2 concentration in wastewater for the dates analyzed. Snowmelt has also been suggested to influence the SARS-CoV-2 viral signal in wastewater (Wade et al., 2022, Statistics Canada, 2022). Although some studies showed the influent volume increased during snowmelt season (Ai et al., 2021, Tiwari et al., 2022), there is a paucity of evidence that snowmelt events have a significant impact on the viral signal.

#### Laboratory factors

Viral concentration measurement from a wastewater sample is a multi-steps process, where each step can introduce a potential source of error. The duration and conditions of transport of the sample from the sampling location to the laboratory may impact the final concentration measurement of SARS-CoV-2. By their nature, wastewater samples are very “active”, i.e. there is a high degree of biological activity that will cause the nature of the sample to change fairly rapidly. The equipment and containers of the sampling system may be contaminated. Therefore, storing, transporting and handling wastewater samples is critical to maintain their integrity and are potential sources of errors. Moreover, the complex and variable nature of wastewater requires to perform controls alongside the molecular detection of SARS-CoV-2 to account for variations in the composition of wastewater and evaluate overall efficiency of the process. Failure to properly run these controls are other potential sources of error. Molecular detection by RT-qPCR may also be prone to potential errors (e.g., standard curve not updated, new viral mutations affecting the identification by primers). Hence, rigorous protocols to ensure consistency and reliability of SARS-CoV-2 concentration measurements from wastewater samples should be in place at this stage of the WBS pipeline. Guidance regarding such protocols are presented in details in Supplementary File 1.

### Normalization

As mentioned above, many factors can perturb the viral concentration in wastewater. Ideally, those factors would be identified, measured, and controlled for before communicating a “final” viral concentration in wastewater. Wastewater is a complex matrix that contains biological, chemical and physical factors that may affect the RNA concentration. It not only collects domestic sewage, but potentially also industrial/agricultural discharges and storm water depending on weather conditions (Nagarkar et al., 2022). From these influents, the composition of wastewater may change in pH, chlorine, and dissolved oxygen contents, which have been suggested to reduce the RNA concentration (Bertels et al., 2022). Moreover, transportation of wastewater through the sewage network involves fluctuations in wastewater temperature, flow rate, sedimentation/resuspension and travel time. For these reasons, it is unlikely to have a consistently smooth viral signal in wastewater, especially when monitoring small communities. However several normalization approaches have been employed by different groups to address these uncertainties in the viral signal. Normalization is not standardized in WBS yet; even the word “normalization” may not be appropriate because it attempts to correct for various factors. Viral signal in wastewater should be controlled for i) human fecal mass to account for population (using biomarkers like Pepper Mild Mottle virus, crAssphage, and ammonia have been suggested); ii) environmental events (e.g., WWTP influent flow), iii) transport and dispersion dynamics in the sewer (e.g., using metrics of particle suspension in wastewater). There is likely no global solution for controlling for these (and other) factors, as each sewer has unique specificities. “Normalization” is still an area of investigation at PHAC/NML, where collection of several normalizing variables (e.g., concentration of Pepper Mild Mottle virus, pH, mass of total solids in suspension) has been performed since the start of the federal WBS program.

### Wastewater Epidemiologic Model

A mathematical model that describes both SARS-CoV-2 transmission at the population level and SARS-CoV-2 concentration in the wastewater (by explicitly modelling fecal shedding) was developed at PHAC/NML (Nourbakhsh et al., 2022) and implemented as a publicly available R package (https://github.com/phac-nml-phrsd/wem). A simple representation of this model, called WEM for wastewater epidemic model, is shown in Figure 3.

**FIGURE 3.**
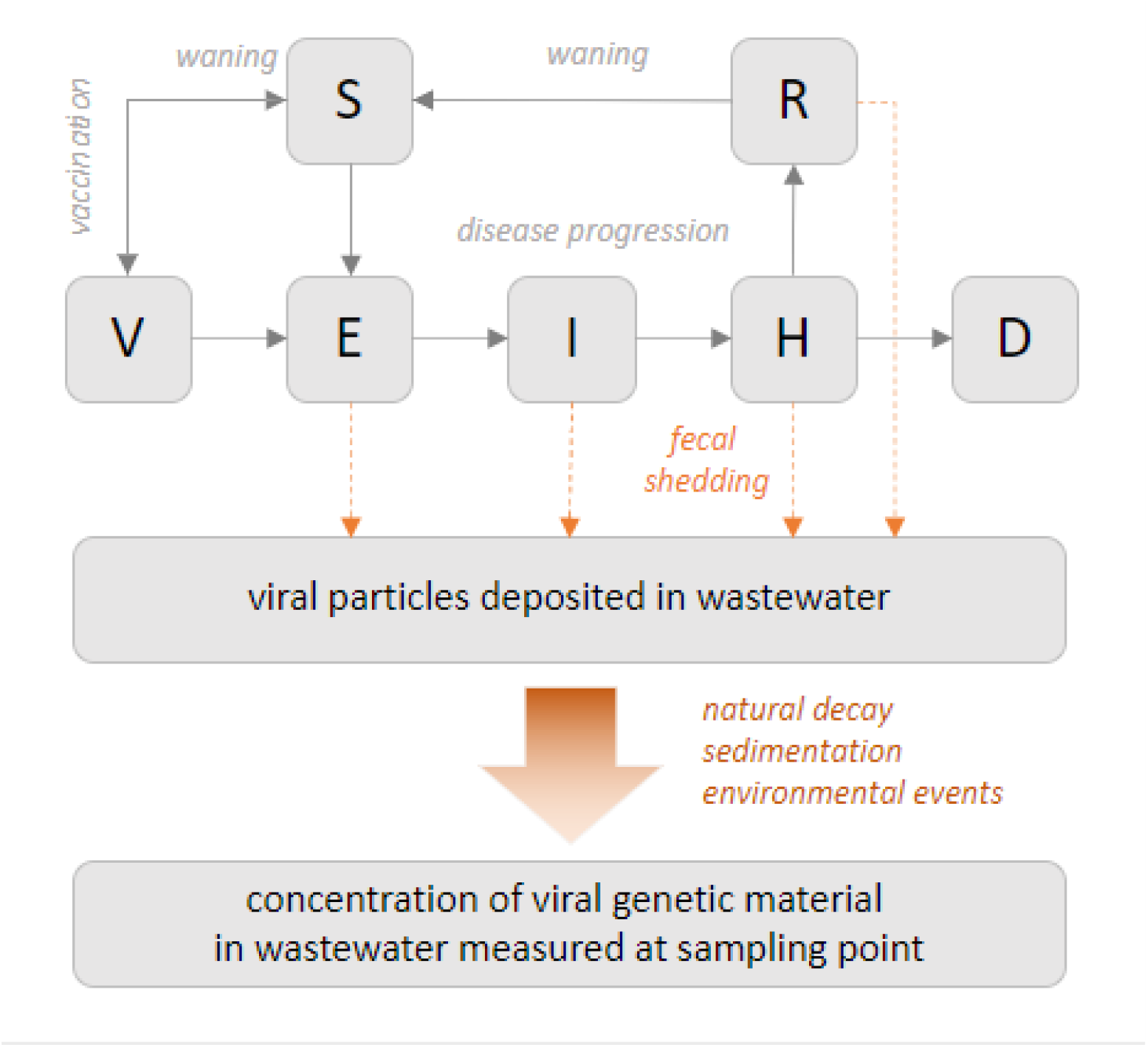
The model is based on standard mathematical modelling of disease spread in population (“SEIR-like”). Different stages of disease progression are represented: susceptible (S), latent exposure (E), infectious (I), hospitalized (H), recovered (R), deceased (D). Immunization is also modelled (V) as well as waning immunity (natural and vaccine-induced). Fecal shedding is explicitly modelled, with the shedding dynamics informed by clinical studies. The viral particles shed in wastewater travel through sewershed up to the sampling point. The model has a simplistic representation of this journey, to account for the effects that can impact the measured concentration, caused for example by the decay of the genetic material of the virus, particles sedimentation and recirculation, and potential impact of environmental events (e.g., large rainfall). The model is calibrated, simultaneously or independently, on clinical data (e.g., reported cases, hospitalizations) and wastewater data (viral concentration).

Like other mathematical models, WEM provides a principled framework to estimate unobserved epidemiological parameters (e.g., actual prevalence, effective reproduction number R_t_) and to forecast cases, hospitalization, and deaths. Importantly, WEM incorporates the wastewater data in addition to the traditional data based on clinical surveillance. The two data types, wastewater and clinical, can be used either in combination when more information is needed to triangulate the state of the pandemic, or as a substitute for one another when one of the two data source is missing. We provide an example of the latter in the section analyzing the Omicron wave.

Because WEM integrates wastewater data, it translates the wastewater signal – that can be hard to interpret epidemiologically – into practical and well-known metrics for public health. Our default approach is to simply use raw (unnormalized) SARS-CoV-2 concentration in wastewater since normalization is still an area of investigation at NML. The case forecasts are key indicators in planning public health actions because they predict the transmission of disease at the population level. The effective reproduction number (R_t_) is another important measure that summarizes the current state of transmission dynamics. These epidemiological indicators of virus transmission played an important role in the national COVID-19 surveillance, and modelling allows to incorporate information from WBS to enhance the estimation of these indicators.

### Wastewater-based surveillance reporting

WBS is, by nature, conducted locally, typically at the level of a municipality (sampling at a WWTP), a neighborhood (sampling in a manhole), or an institution (e.g., hospital, university campus). When data from several sampling sites are available, it may be more relevant to aggregate the data to provide a trend indicator for a broader geographical area. A naïve approach to aggregate viral concentrations in wastewater from different sites is to perform a weighted average where the weights represent the population sizes of each catchment area. Of course, the viral concentrations must be standardized beforehand.

To inform its analyses, PHAC, a national agency, aggregates WBS from samples collected at the wastewater treatment plants to municipal and national levels. PHAC analyzes WBS through the lens of modelling. Hence, the weighted average aggregation is performed on the epidemiological metrics (e.g., forecasted incidence, R_t_) after fitting WEM to the data of each sampling sites. In other words, we do not fit WEM to an aggregated wastewater signal. WBS is reported in combination with clinical surveillance and modelling forecasts to show the wastewater concentration and cases to date, and predictions based on WEM.

### Application to the analysis of the Omicron wave in Canada

The Omicron variant of SARS-CoV-2 was classified as a VOC on Nov 26, 2021 (World Health Organization, 2021). By January 2022, over 90% of SARS-CoV-2 samples collected in Canada were identified as Omicron (Ontario Agency for Health Protection and Promotion (Public Health Ontario), 2022). Omicron spread rapidly across Canada, which prompted a change in testing policies to restrict PCR testing to high-risk or vulnerable populations in many jurisdictions to meet the overwhelming demand. This change likely led to an underestimation of disease burden by clinical surveillance. Importantly, case forecasts from models using the case data could no longer serve as reliable indicators to inform public health policies. In fact, in all five cities analyzed with WEM, wastewater viral loads increased concordantly with clinical cases, but the trends diverged with the implementation of PCR testing restrictions. While clinical cases appeared to have peaked around the date of the restriction, wastewater signals continued to increase or remained elevated. The discordance between clinical surveillance and WBS during the Omicron wave emphasized the utility of WBS when clinical testing was restricted (Statistics Canada, 2022).

To provide more accurate case estimates and forecasting with WEM in absence of reliable clinical testing, the model was calibrated alternatively to clinical and WBS data. In addition, model parameters such as asymptomatic proportion and vaccine efficacy were re-defined to account for Omicron-specific transmission dynamics. After these adjustments were made, data from WBS, clinical surveillance and model forecasts were reported with epidemiological interpretations for internal monitoring of the national SARS-CoV-2 trends (Figure 1). WEM provided estimates of reportable cases (i.e., clinical cases that would have been reported without PCR testing restrictions) using wastewater data only, in comparison with actual reported clinical cases, to assess the extent of under-reporting and the likelihood of having passed peak incidence of the wave. In Figure 4, we illustrate how modelling outputs were used in two different cities during the Omicron wave. In this example, WEM was fit alternatively to clinical or wastewater data in Toronto (the largest city in Canada) and Edmonton (a medium size city). The model suggests that under-reporting of cases in the former was more pronounced than in the latter. This modelling analysis of the Omicron wave highlighted the limitations of clinical surveillance, especially after the change in PCR testing guidelines. From WBS, the under-reporting of cases was evident through the comparison of cases estimated from clinical surveillance and WBS. Moreover, WBS complemented the information from clinical surveillance including the timing of the peak and increasing/decreasing trends. Overall, the Omicron wave in Canada has allowed for an appreciation for the utility of WBS as an alternative approach to monitor SARS-CoV-2 transmission when clinical surveillance became overwhelmed and struggled to provide high quality data on disease prevalence trends.

**FIGURE 4.**
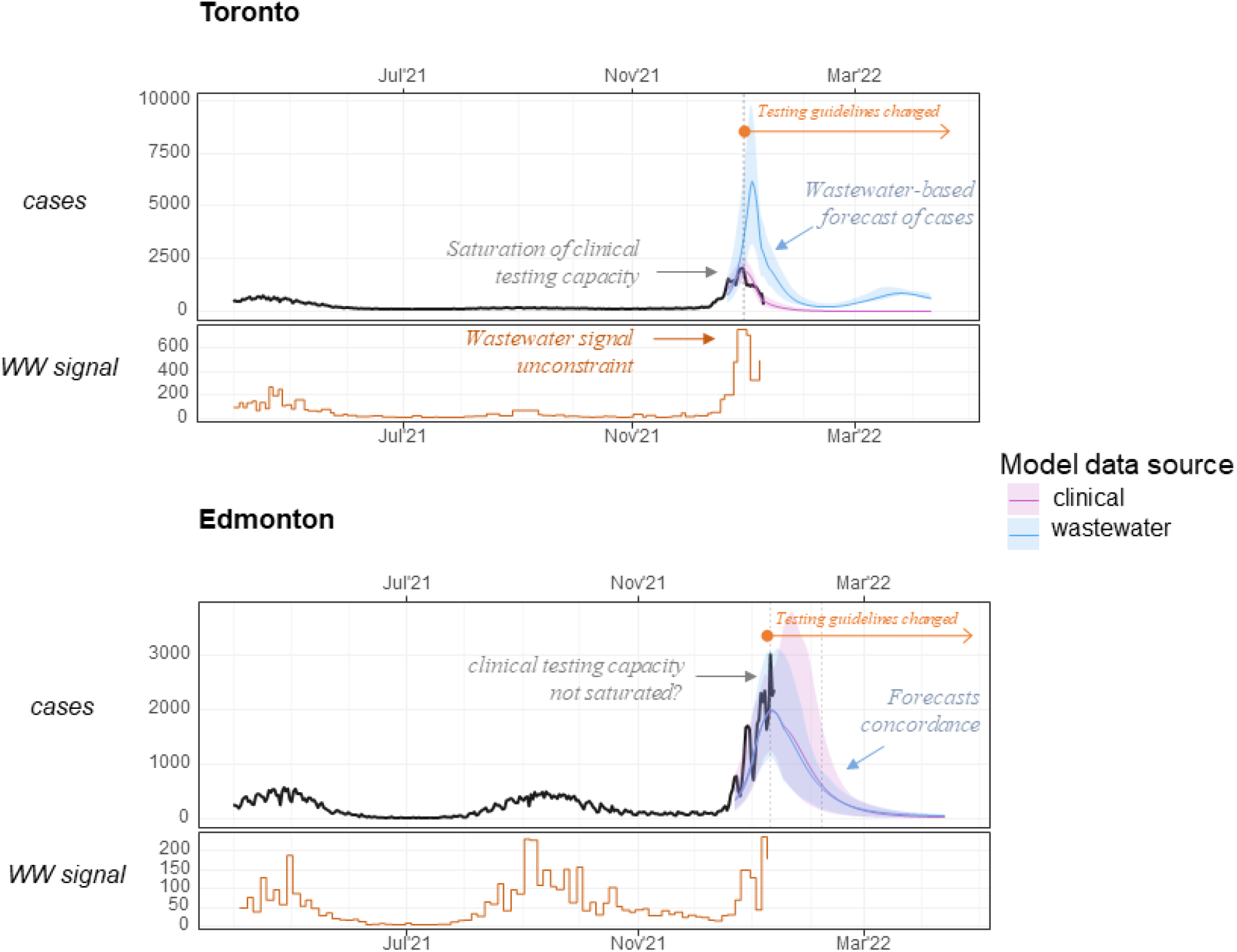
Example of model output interpretation during the Omicron wave. For each city, the op panel shows the reported clinical cases (black curve) and the model-inferred reportable cases using clinical reports (pink curve) or using the wastewater signal (blue curve). The bottom panel shows the average SARS-CoV-2 concentration in wastewater in the city (orange step line). The vertical dotted line indicates when the municipal testing guidelines were restricted. The difference between the blue and pink curve is proportional to the extent of under-reporting of clinical cases. In Toronto, modelling suggests that under-reporting was significant around peak incidence, whereas Edmonton seems to not have been affected as much. Analysis performed as of 2022-01-19.

### Limitations of wastewater-based surveillance in Canada

Currently, the wastewater-based modelling focuses on five major cities in Canada. While the combined catchment area of WBS for these five cities is about 23% of the Canadian population (Statistic Canada 2021 census, https://www12.statcan.gc.ca/census-recensement/2021/dp-pd/prof/index.cfm?Lang=E), it cannot provide a comprehensive overview of the SARS-CoV-2 trends with the limited scope of surveillance. Although the expansion of the CWS may increase the coverage of WBS, several challenges are anticipated given the geography and population distributions in Canada.

First, WBS in small or remote communities will require different sampling methods, such as sample collection from a septic tank, manhole, or lagoon, due to the absence of WWTPs in such areas. Although previous research has demonstrated that sampling from manholes did not result in significant RNA decay (Yeager et al., 2021), it poses as a logistical challenge. Moreover, our current modelling framework (WEM) is not adapted to analyze small populations, mainly because WEM is not a stochastic model.

Although still an area of active research, controlling for uncertainty in the viral signal in wastewater, such as fecal shedding dynamics and in-sewer RNA decay, is critical. Since the viral signal is meant to be used to inform public health, normalization should improve its specificity and sensitivity. The uncertainty of normalization techniques, at PHAC/NML but also for many other groups, is currently a limitation when interpreting WBS.

### Beyond COVID-19

The implementation of WBS as a routine surveillance tool has broader implications beyond COVID-19. WBS can also be used for monitoring other respiratory pathogens other than SARS-CoV-2 such as influenza viruses or respiratory syncytial virus, sexually transmitted infections, antibiotic resistance, and antibiotic use in the community (Heijnen & Medema, 2011; Zhang et al., 2019). Importantly, the active research of WBS during the COVID-19 pandemic allowed for a better understanding of in-sewer factors, environmental factors and population dynamics that affect WBS and the development of mathematical modelling to estimate population prevalence of the health risk and its future predictions. However, we note that for any pathogen surveilled in wastewater, it is critical to understand its fecal shedding dynamics and in-sewer decay to improve estimates of infection prevalence in the community from viral concentration measured in wastewater. Unfortunately, there is a dearth of such clinical studies, even for SARS-CoV-2. While the expansion of WBS to other pathologies will require the development of novel laboratory assays, the current framework and knowledge of WBS and modelling with WEM will provide a strong foundation to facilitate the surveillance of other infectious disease pathogens.

### Next steps

While the present framework provides a comprehensive analysis pipeline for the current scope of national WBS, changes and improvements can be implemented to respond to the dynamic nature of the COVID-19 pandemic. A crucial step in further developing WBS is to standardize the surveillance data, including its measurement metrics and storage, across many laboratories. The Public Health Environmental Surveillance Open Data Model (PHES-ODM, https://github.com/Big-Life-Lab/PHES-ODM) is an initiative to develop an open data structure, including metadata and vocabulary, to support environmental surveillance such as WBS. PHAC is in the process of incorporating its national WBS into the PHES-ODM to augment its capacity to monitor multiple pathogens and geographical locations for WBS (facilitating the scalability of data analysis, thanks to its standardized data structure). In addition to incorporating data from concurrent WBS programs, WBS has the potential to expand to more geographical locations with diverse environments, such as remote or small communities. However, remote communities pose unique challenges because they often lack a WWTP and require alternative sampling methods for WBS. Hence, the framework may also expand to incorporate data analysis processes from these varying sources of WBS samples to standardize the analyses. Lastly, WBS can serve as an indicator of emerging VOCs through SARS-CoV-2’s genome sequencing. Although this is currently conducted at the NML, the epidemiological interpretations of the results are not yet incorporated in the pipeline described here.

## Conclusion

Although WBS has previously been used to inform public health responses for other health risks, the COVID-19 pandemic incited an expansion of WBS to an unprecedented scale. As demonstrated during the Omicron wave, COVID-19 WBS has the potential to have high policy implications, especially when traditional epidemiological surveillance methods curtail. The present framework outlines the analysis pipeline of the first national WBS of COVID-19 in Canada. In particular, the use of mathematical modelling is a critical tool to interpret WBS. While WBS of COVID-19 provides unique information on the community spread of SARS-CoV-2, there still remain many uncertainties and inconsistencies to be explained in the WBS data. The establishment of this framework will support further expansions and development of the WBS program, such as monitoring for other geographical areas and other pathogens in wastewater.

## Data Availability

All data produced in the present study are available upon reasonable request to the authors

## List of Abbreviations

(CS): Combined sewer;
(CWS): Canadian Wastewater Survey;
(COVID-19): Coronavirus Disease 2019;
(R_t_): Effective reproduction number;
(NML): National Microbiology Laboratory;
(PHAC): Public Health Agency of Canada;
(WBS): Wastewater-based Surveillance;
(WWTP): Wastewater treatment plant

