## Supplementary file S1 for "Coupling Wastewater-Based Epidemiological Surveillance and Modelling of SARS-COV-2/COVID-19: Practical Applications at the Public Health Agency of Canada"

### Coupling Wastewater-Based Epidemiological Surveillance and Modelling of SARS-COV-2/COVID-19: Practical Applications at the Public Health Agency of Canada

Meong Jin Joung[1,4], Chand S Mangat[2], Edgard Mejia[2], Audra Nagasawa[3], Anil Nichani[2], Carol Perez-Iratxeta[3], Shelley W Peterson[2], David Champredon[1,*]

[1] Public Health Agency of Canada, National Microbiology Laboratory, Public Health Risk Sciences Division, Guelph, ON, Canada.

[2] Public Health Agency of Canada, National Microbiology Laboratory, One Health Division, Winnipeg, MB, Canada.

[3] Statistics Canada, Ottawa, ON, Canada.

[4] Dalla Lana School of Public Health, University of Toronto. Toronto, ON, Canada.

#### Laboratory analysis of SARS-CoV-2 concentration

##### *Sampling Frequency*

As most wastewater surveillance activities are limited, daily sampling is often not possible. Typically, labs collect 2 to 3 samples per week which are often sent in-batch to a testing laboratory. To most accurately describe the trends of SARS-CoV-2 concentration in wastewater, these samples should be collected in well-spaced intervals throughout the week.

##### *Sample Handling, Transportation, Storage, and Quality Control*

By their nature, wastewater samples are very “active”, i.e. there is a high degree of biological activity that will cause the nature of the sample to change fairly rapidly. After collection, samples should be cooled to 4°C as quickly as possible, then shipped cold using natural ice or ice packs via the most rapid available transportation. Upon arrival at the laboratory, they should be extracted as soon as possible, keeping the samples refrigerated until extraction. Standard Methods[1] recommends extraction within 7 days of collection when analyzing for trace contaminants such as semi-volatile organics. Wastewater samples also contain suspended solids, which are an integral part of the matrix. As such, samples must be shaken frequently and thoroughly during any sub-sampling in the field or laboratory.

Composite and grab sampling includes the use of consumables (tubing, bottles) and reusable containers and equipment. These containers and equipment must be tested to ensure that the sampling system is not introducing contaminants into the samples. Laboratory-grade water can be used to create Equipment Blanks by simulating a composite or grab sampling event that includes sample tubing, pump tubing, collection containers and sub-sampling containers.

#### Wastewater Treatment Plan Metadata and Context

As discussed above, a wastewater study must be designed in the context of the collection and treatment system realities and details. Wastewater samples should always be characterized for conventional parameters to provide the context of wastewater strength and effectiveness of the treatment process. These parameters are listed in Table S1.

#### Summary

Any study of wastewater constituents requires a thorough understanding of the collection and treatment system, in order to design a sample collection process that will answer the study questions. Sampling locations should be confirmed in consultation with WWTP operators, and described in details in all reports and publications (for example: collection of raw influent or after screening process). Likewise, sampling techniques (composite or grab) should be described in sufficient detail. Wastewater samples must be stored, transported, and handled appropriately to maintain their integrity.

*Table S1: Conventional wastewater parameters (APHA et al 2017)[1]*

| Parameter | Comments |
|---|---|
| **Temperature - process** | Indicator of microbial conditions for treatment |
| **Temperature - sample** | Confirmation of target sampling temperature |
| **pH** | Indicator for general chemistry and microbiology |
| **Alkalinity** | Indicator of buffering capacity and nitrification |
| **Total Suspended Solids (TSS)** | Empirical gravimetric test, indicator of wastewater strength and treatment effectiveness, can be correlated with some chemical and microbiological constituents |
| **Chemical Oxygen Demand (COD)** | Measure of material amenable to oxidation under strong chemical conditions, indicator of wastewater strength and treatment effectiveness |
| **Biochemical Oxygen Demand (BOD)** | Measure of material amenable to oxidation under specific biological conditions, indicator of wastewater strength and treatment effectiveness |
| **Total Organic Carbon (TOC)** | Measure of total organic (reduced) carbon, indicator of wastewater strength and treatment effectiveness |
| **Total Kjeldahl Nitrogen (TKN)** | Measure of total organic (reduced) nitrogen |
| **Ammonia nitrogen** | Measure of nitrogen available for nitrification |
| **Nitrate + nitrite** | Measure of oxidized nitrogen, indicator of nitrification or denitrification |
| **Measured average daily flow** | Available from the WWTP, indicates the size of the system and confirms dry weather conditions or influence of storm events |

#### Laboratory processing of wastewater sample for SARS-CoV-2 RNA detection

Once samples are received at the lab, the temperature should be taken and recorded for future reference. Samples should be processed within 24 hours of receiving them in the lab. Effective wastewater surveillance that aims to detect the emergence of infection relies on rapid data collection and testing. For future use, store unused portions/aliquots of collected samples at -70°C. Since the strength of the viral RNA signal decreases after freezing, consequently more than one freeze-thaw cycle should be avoided. Figure X describes the laboratory processing steps employed to detect a SARS-CoV-2 RNA signal from a wastewater sample.

***Figure S1:*** *Laboratory Processing of Wastewater sample for identification of SARS-CoV-2*

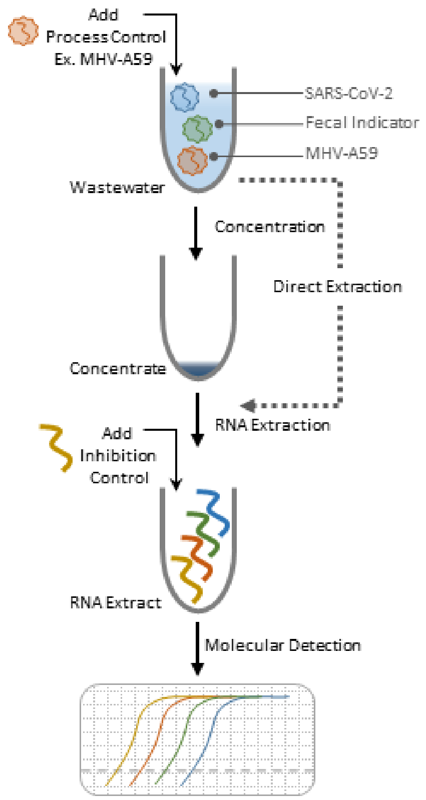


#### *Laboratory testing Procedures*

A wastewater test for SARS-CoV-2 is comprised of three major steps (Figure 5):

(i) Viral concentration,

(ii) RNA extraction, and

(iii) molecular detection.

Currently, the PHAC-NML employs a variant of an ultrafiltration-based viral concentration method previously described by the Wigginton group[2] in addition to an assay directed at the "solids" or insoluble fraction of wastewater followed by molecular detection of SARS-CoV-2 by RT-qPCR directed at the N1 and N2 targets as developed by the US-CDC. Previous studies have shown that SARS-CoV-2 is roughly partitioned equally between the solid and liquid phase of wastewater. Volumetrically, the solids fraction is the major component of wastewater that holds the virus[3-6]. The solids fraction can be easily collected and extracted to yield genetic material suitable for testing.

There is no consensus or gold standard test for SARS-CoV-2 detection in wastewater and inter-laboratory comparisons of methods have shown that most perform comparably. In collaboration with the Canadian Water Network, PHAC-NML participated in an inter-laboratory study of SAR-CoV-2 wastewater detection methods[7]. The sample was drawn from the largest of three Winnipeg WWTP and there were 85 clinical cases across the city at the time of collection. SARS-CoV-2 concentrations from most labs were within a 1-log band of each other. The Water Research Foundation[8] performed a similar inter-laboratory comparison amongst US laboratories. Grab samples from two WWTPs servicing Los Angeles County (~ 30K cases reported in the previous 14 days) were distributed to 36 laboratories for analysis. Despite methodological differences, range of reported results from most laboratories were within a 2-log band. Importantly, the above studies show that there was no consensus in method amongst participating laboratories, suggesting that most methods performed comparably.

The structure of the Canadian study revealed important considerations for laboratory methods. Eight laboratories received three sample types; samples spiked with inactivated SARS-CoV-2 at a high and low concentration (1,800 cp/mL vs. 20 cp/mL), and an unspiked sample. First, only laboratories that processed the insoluble or “solids” fraction of wastewater were able to derive signal from the unspiked sample. Because of the low clinical case count at the time of collection, this suggests that the insoluble fraction of wastewater is more effective for delivering early-warning indicators from wastewater surveillance. Studies of primary sludge and fractionation of wastewater influent have confirmed that the majority of the SARS-CoV-2 viral signal resides in the insoluble fraction. Secondly, the SARS-CoV-2 spiked into wastewater did not appreciably partition to the insoluble or “solids” fraction of wastewater. This suggests that the use of surrogate virus controls to monitor the overall efficiency of laboratory methods may not report on natural viral signal (See Controls, below).

*Laboratory Methods*

#### Viral concentration

As SARS-CoV-2 is found at low levels within wastewater, concentration is required for accurate analysis, especially during the initial outbreak phases when the viral load is low (Figure S1). Concentration and extraction are widely considered as the most influential steps in directing the overall performance of the assay. Ahmed *et al.* have elaborated comparatively on the different methods[9]. There are a variety of concentration methods, each with their own advantages and disadvantages as described in Table S2.

Some investigators employ direct extraction schemes on whole wastewater that skip the concentration step altogether (Figure S1). Typically, this involves processing about 1 mL of whole influent, which is both practical and amenable to high-throughput. Direct extraction avoids sample losses associated with concentration and could improve overall yield. However, PHAC-NML cautions against using direct extraction schemes because of their unknown performance during periods of low viral load, especially when testing low volumes of wastewater.

*Table S2. Three common concentration methods*

| Method | Advantages | Disadvantages |
|---|---|---|
| • **Ultrafiltration**<br>• **Viral particles are concentrated by the use of centrifugal filter device.** | • Easy to use<br>• Short turn-around time<br>• Higher-throughput than most methods<br>• Doesn't access solids component | • Co-concentrations of inhibitory compounds<br>• Cost of labware (~$30 per device)<br>• Supply of reagent may vary<br>• Requires centrifuge (to 4K x g)<br>• Filters can clog when sample turbidity is high |
| • **Electronegative filtration**<br>• **Viral particles are captured on a charged membrane by vacuum filtration** | • Low Cost<br>• Low carryover of inhibitory compounds<br>• Low lab overhead to install test | • Low-throughput<br>• High-hands on time<br>• Requires extensive RNA extraction/clean-up |
| • **PEG precipitation**<br>• **A precipitating agent is added to samples and viral particles are recovered by centrifugation** | • Low Cost<br>• High supply of reagents | • Low throughput, some methods require overnight incubation<br>• Requires centrifugation (to 10K x g)<br>• Requires extensive RNA extraction/clean-up |

As discussed above, the majority of the viral signal is associated with the insoluble fraction of wastewater. As such, collection and processing of wastewater solids could improve recovery dramatically. PHAC-NML has found that the viral level in wastewater solids is equal to, or greater than, the liquids fraction.

##### *RNA extraction*

There are a variety of commercial RNA extraction kits available and each should be chosen based on the type on input material. Samples with high-solids content require mechanical disruption and extensive wash steps to remove inhibitory compounds; "soil" or "microbiome" extraction kits are well suited for this purpose. General RNA extraction kits can be employed when the input material is clarified by centrifugation. Commercial clean-up kits can improve detection in inhibited samples. RNA is unstable once extracted, therefore molecular detection should be performed the day of extraction.

##### *Molecular Detection*

Detection of the viral signal from SARS-CoV-2 is by reverse transcription quantitative polymerase chain reaction (RT-qPCR) as indicated in Figure X. Specific primers amplify the SARS-CoV-2 genome and an intervening fluorescent probe is concomitantly consumed in this process. The viral signal is monitored by the increase in fluorescence associated with the consumption of this probe. Quantitation is achieved by measuring the number of cycles (Cycle threshold or Ct) required for the fluorescence detection of consumed probe over a baseline value "threshold", which is compared to a standard curve of known input quantities. A consistent "threshold" value should be used for all samples and the "auto-thresholding" function of the RT-qPCR instrument should be disabled. The threshold is specific to the RT-qPCR instrument and primers/probes chosen for analysis require optimization to reduce noise between replicates.

Sequences for SARS-CoV-2 primers, probes, controls, and sequences required for quantification above are provided in appendix 2. There are several established primer/probe sets used to detect SARS-CoV-2 in wastewater and there is no current consensus as to which molecular targets are best. Indeed, investigators have reported contradictory performances of the same primer/probe combinations. PHAC-NML has evaluated the *E-Sarbeco*[10] and US-CDC N1/N2 targets and found N1/N2 to be the most sensitive and consistent. The NML recommends using two targets to mitigate the risk of mutation.

##### *Controls*

The complex and variable nature of wastewater requires three controls run alongside the molecular detection of SARS-CoV-2 to account for variations in the composition of wastewater and evaluate overall efficiency of the process.

#### Process control

To account for varying efficiencies of the RNA extraction from wastewater a spike-in control of whole-viral material is added to wastewater prior to concentration as described in Figure 3. A parallel concentration/extraction is run in PBS and the overall concentration and extraction efficiency is inferred by comparing the relative recovery. The process control or surrogate is ideally a coronavirus of the same genus as SARS-CoV-2 and thus physically structured similarly to SARS-CoV-2 to best report on its recovery. Common process controls are murine hepatitis virus (MHV-A59), bovine coronavirus or one of the seasonal human coronaviruses. PHAC-NML currently adds Mouse Hepatitis Virus A59 (MHV-A59) as a process control in its assays. PHAC-NML has found that cultured MHV-A59 does not appreciably partition to the solids phase of wastewater and thus is not reflective of the natural state of the virus (as discussed above). A similar lack of solids-phase partitioning has been observed from cell-culture-produced SARS-CoV-2. Therefore, the utility of surrogates is likely more suited to methods that process only the liquid fraction of wastewater.

#### Fecal control

The fecal load of wastewater can vary across wastewater collection systems. Surface water, ground water, and varying industrial and institutional inputs can dilute wastewater and introduce variance to the SARS-CoV-2 signal. A test specific for fecal load is applied to account for the varying composition of wastewater. PHAC-NML currently directs a RT-qPCR reaction against the Pepper Mild Mottle Virus (PMMoV), a naturally occurring virus that is found abundantly in edible peppers and reports on fecal load[11]. Other fecal indicators of note are the HF183 and crAssphage.[12, 13]

#### Inhibition control

Wastewater contains contaminants that are known to inhibit PCR assays. To detect the presence of inhibitors, purified RNA from a source that is not found in wastewater is added to wastewater RNA extracts or alternatively to the wastewater concentrates. The signal intensity of this reaction is compared to the inhibition indicator material tested alone. The Water Research Foundation inter-laboratory study suggests that a shift of ≥1 Ct suggests absence of PCR inhibition[8]. An alternative approach when purified RNA is not available is to dilute a wastewater RT-qPCR reaction and compare the resultant Ct-value with the expected value[14]. When inhibition is outside of one Ct of the expected range, it is suggested to dilute the wastewater prior to extraction and/or to flag the results prior to reporting.

##### Negative controls

Good practice with RT-qPCR based experiments is to run a RT-qPCR reaction without the addition of template. Any signal observed in this control would indicate the presence of contaminants in the RT-qPCR reagents. Mock concentration/extractions using buffer or water alone should be run periodically to identify contaminated labware or reagents.

##### *Normalizing Techniques*

Following quantification, the SARS-CoV-2 signal and associated controls are expressed in copies per volume of the processed wastewater (e.g. cp/mL). Adjustments should be made for wastewater losses over concentration (e.g. unrecoverable dead volume in centrifugal filter devices) and/or dilution of samples prior to extraction (e.g. to mitigate inhibition if observed). Estimation of the dead-volume can be made by determining the weight difference of the centrifugal filter device before the application of sample and after sample recovery, assuming a density of 1 g/mL.

Quantified viral targets are normalized to the quantified fecal indicator and this value alongside the un-normalized data should be considered minimal for reporting (Eq 1.).

$$\frac{Viral\ target\ (\frac{copies}{mL})}{Fecal\ Indicator\ (\frac{copies}{mL})} = Fecal\ normalized\ (copies/mL)\ \text{(Eq. 1)}$$

Further adjustments to the reported value can be made by incorporating the yield of the process control to overall yield. First, overall yield is calculated (Eq.2)

$$\frac{Process\ control\ copies\ recovered}{Process\ control\ copies\ in} = Process\ control\ yield \text{ (Eq. 2)}$$

To adjust for yield process recovery, apply the following formula to calculate the yield adjusted viral load (Eq 3.)

$$\frac{1}{Process\ control\ yield} * Fecal\ normalized\ \left(\frac{copies}{mL}\right) = Yield\ adjusted\ viral\ load$$

As described above, the process control may not accurately report on the overall SARS-CoV-2 yield, especially where the solids fraction is primary target for extraction. Where possible, flow-based normalization is the preferred method to account for dilution effects, which can be a major contributor to the loss of signal - particularly during wet weather events.

14. King CE, Debruyne R, Kuch M, Shwarz C, and Poinar HN. A quantitative approach to detect and overcome PCR inhibition in ancient DNA extracts. BioTechniques 5 (47). doi: https://doi.org/10.2144/000113244.
